# The role of veterinary diagnostic laboratories during COVID-19 response in the United States

**DOI:** 10.1101/2023.12.05.23299493

**Authors:** Nia Clements, Diego G. Diel, François Elvinger, Gary Koretzky, Julie Siler, Lorin D. Warnick

## Abstract

Robust testing capacity was necessary for public health agencies to respond to severe acute respiratory syndrome coronavirus 2 (SARS-CoV-2) during the coronavirus disease 19 (COVID-19) pandemic. As the nation faced the need for robust testing capacity, it became necessary to use all possible resources. In many cases, veterinary diagnostic laboratories rose to meet this demand because these facilities routinely perform high throughput diagnostic testing of large animal populations and are typically familiar with pathogens of high pandemic concern. In this study, we evaluated the impact of veterinary diagnostic laboratories in the United States on SARS-CoV-2 testing. Results of surveys, semi-structured interviews, and analysis of publicly available information showed that veterinary diagnostic laboratories had a substantial impact on human health through population-level testing in the COVID-19 response, supporting timely and informed public health interventions. This success was not without significant hurdles, as many participating veterinary diagnostic laboratories experienced restriction in their response due to difficulties obtaining the Clinical Laboratory Improvement Amendments (CLIA) certification required to conduct human diagnostic testing. Our results point out the importance of reducing hurdles before the next major public health emergency to enhance access to testing resources overall and to ultimately improve population health.

## Introduction

Severe acute respiratory syndrome coronavirus-2 (SARS-CoV-2) was first detected in Wuhan, Hubei Province, China when numerous cases of atypical pneumonia linked to the Hunan Seafood Market emerged in December of 2019.[1,2] Soon after the initial cases in China, SARS-CoV-2 was detected and reported in the United States (US) on January 21, 2020 in Washington State in a patient with respiratory illness.[2] This case was determined to be epidemiologically linked to Wuhan, China.[2] As clinical case numbers mounted, a public health emergency was declared 10 days later on January 31 by the United States Department of Health and Human Services.[2] By this time, the disease was rapidly spreading worldwide, triggering mass shutdowns to protect public health while key characteristics of the disease remained unknown.

The need for robust SARS-CoV-2 testing was apparent from the pandemic’s start. When the outbreak was in its early stages, testing was concentrated in US major ports of entry to mitigate travel-related spread.[2] The need to expand testing became urgent with evidence for local spread. Additionally, key epidemiological characteristics of the virus were revealed, such as SARS-CoV-2’s early basic reproduction number between 2.2 and 2.7[3] and the high incidence of asymptomatic disease and efficient virus transmission.[1] The emergence of non-travel related cases, coupled with the epidemiological characteristics of SARS-CoV-2, sounded an alarm to the highly infectious nature of the virus and made it clear that broad and aggressive testing would be required to control virus spread. Public health messaging emphasized this point, communicating that testing was necessary to “flatten the epidemic curve,” referring to a strategy of reducing the number of new disease cases by using public health interventions, including testing, so that those infected would be aware and take appropriate measures to prevent viral transmission.[4,5] Countries that incorporated a rigorous testing approach as part of their strategy to “flatten the curve” experienced relative success in controlling and decreasing the spread of the virus. For example, with a population of 51.8 million,[6] South Korea achieved a SARS-CoV-2 testing capacity of 15,000 tests per day early in the pandemic.[4] High testing capacity was achieved through accessible drive-through and walk-in testing sites, allowing for a testing rate of 17,000 tests per million people by June 2020.[4] This robust testing approach was a factor among other strategies (e.g. extensive contact tracing, rigid quarantine rules)[7] that prevented the need for lockdowns. South Korea’s public health solution to SARS-CoV-2 consequently had numerous positive downstream effects, such as maintenance of livelihoods in communities and diminishing economic disruption.[4] Given South Korea’s SARS-CoV-2 outcomes, it is clear that effective control was achievable, emphasizing the importance of high testing capacity, among other strategies, in this emergency response.

One approach to achieve high testing capacity is through the collaboration of human diagnostic laboratories and veterinary diagnostic laboratories. As stated by the World Organization for Animal Health, veterinary diagnostic laboratories are well suited to supporting testing responses in public health emergencies “because they have experience in quality assurance, biosafety and biosecurity, and high throughput testing for the surveillance and control of infectious diseases in animals, some of which are zoonotic.”[8] Importantly for quality control, veterinary diagnostic laboratories, like human health laboratories, have external accreditors that ensure testing is performed under guidelines that promote accuracy and workforce competence[9] and have Biological Safety Level 2 and 3 facilities to handle diseases of public and animal health concern.[10]

In the throes of a public health crisis due to zoonotic infection, veterinary and animal diagnostic laboratories have qualities that may make them more nimble than human health laboratories. For instance, veterinary diagnostic laboratories strive to develop novel testing programs given their familiarity with a wide range of pathogens that have zoonotic potential.[11] Furthermore, animal laboratories have extensive experience in surveilling livestock and poultry for highly pathogenic diseases as threats emerge.[12] Because veterinary diagnostic facilities surveille entire populations of animals to protect the agriculture industry, these laboratories are designed to conduct “herd” testing, whereas human diagnostic laboratories focus on testing individuals. This testing capacity of veterinary laboratories provides a unique opportunity to enable entire populations of humans to be tested in the face of health threats like SARS-CoV-2. Given these characteristics, a collaboration between human health and veterinary diagnostic laboratories can synergize to develop testing programs during pandemics.

This potential for collaboration between human diagnostic laboratories and animal diagnostic laboratories is already recognized from a regulatory standpoint, as human-focused diagnostic initiatives can be housed in veterinary diagnostic laboratories with Clinical Laboratory Improvement Amendments (CLIA) certification by the Centers for Medicare and Medicaid Services.[13] For instance, CLIA enabled seven of the sixty member laboratories of the National Animal Health Laboratory Network which is coordinated by the USDA Animal and Plant Health Inspection Service to be certified for human SARS-CoV-2 samples by June 2020, early in the pandemic.[14]

The objective of this study was to determine the extent and quantify the impact of SARS-CoV-2 testing in veterinary diagnostic laboratories to better understand their role in the SARS-CoV-2 response, and to inform decisions for future public health emergencies among veterinary laboratories, human health organizations, and regulatory bodies such as the Centers for Medicare and Medicaid Services.

## Materials and methods

### Survey of veterinary diagnostic laboratories

#### Source population

Following formal survey approval by the American Association of Veterinary Medical Colleges, an online cross-sectional Qualtrics survey was distributed to American Association of Veterinary Medical Colleges and the National Animal Health Laboratory Network member laboratories in the United States using email listservs. If publicly available information detailed that a laboratory had a robust SARS-CoV-2 testing program but no response was recorded for this laboratory following the survey’s due date, laboratory leadership were contacted directly via email to personally invite them to participate. Responses were collected between March 31, 2023 and July 18, 2023.

#### Survey

Respondents first identified the name of their laboratory and whether they conducted animal SARS-CoV-2 diagnostic testing and human SARS-CoV-2 testing. Respondents who conducted animal SARS-CoV-2 testing were asked 6 questions about the duration of the testing program, number of animals tested, testing methodology, and if viral nucleic acid sequencing was conducted. Respondents whose laboratories conducted human SARS-CoV-2 testing were asked 19 questions across 4 domains, including test characteristics, timeline and testing volumes, testing logistics, and reflections on emergency response. For all questions respondents could either select from an array of response choices or write in a response. At the end of the survey, respondents had the option to include their contact information for follow-up. A copy of the survey may be found in the supporting information (see S1 File).

#### Survey analysis

Responses were analyzed by descriptive statistics and qualitative analysis using Microsoft Excel and R (version 2022.07.2+576 “Spotted Wakerobin”). R packages included readr, questionr, and ggplot2. When curating data for analysis, duplicate laboratory responses were combined into one entry. In the case of numerical response entry (e.g., date of testing commencement, number of total samples tested), the numerical response that was either the most detailed (e.g., August 21, 2023 versus August 2023) or the largest (e.g., 1,245 samples versus 1,000 samples) was used in the final response for laboratories with duplicate responses. Incomplete responses that either could not be traced back to a specific institution or had no meaningful information (e.g., most questions unanswered) were removed from the dataset.

### Semi-structured interviews with survey respondents

#### Source population

To learn more about the specific elements of a laboratory’s response, a convenience sample of survey respondents were asked to participate in a semi-structured interview. Two groups of interviewees were invited: (1) laboratories that conducted human SARS-CoV-2 testing and (2) laboratories that did not conduct human SARS-CoV-2 testing. For the first group, invitations for interview were sent to two survey respondents per region of the US (Northeast, South, Midwest, and West) to include laboratories representing all US regions. Within regions, invitations for interview were selected based on interesting elements of a laboratory’s response (e.g., unusually high testing capacity, testing methodology that differed from what was observed in most responses). Based on the barriers cited by these interview respondents in group 1, laboratories in group 2 were invited for interview to gain a deeper understanding of what barriers may exist in conducting human SARS-CoV-2 testing. One laboratory that did not conduct human SARS-CoV-2 testing per US geographic region was invited to interview, except for the Midwest because all laboratories that responded to the survey in this region conducted human SARS-CoV-2 testing. Across both group 1 and group 2, eleven interview invitations were sent and ten interviews were conducted. One invited laboratory did not respond to requests for interview.

#### Interviews

Interviews lasted approximately 30 minutes and were conducted virtually via Zoom, Microsoft Teams, or by phone. Interviewees were asked about what elements of their testing program were most impactful on their laboratory’s testing capacity, what barriers they experienced in either setting up or operating their testing programs beyond those discussed in the survey responses, and what information they would find useful for their current and future laboratory operations from this publication. If time permitted, interviewees were asked for their major learning outcomes and anticipated changes to pandemic preparedness following their experiences with SARS-CoV-2. For interviewees whose institutions did not conduct SARS-CoV-2 testing, additional questions were asked about their laboratory’s regular operations and if there was consideration of using veterinary laboratory resources during their institution’s COVID-19 response. A copy of interview questions may be found in the supporting information (see S2 File).

#### Interview analysis

During all but one interview, two researchers were present for interview facilitation and note taking. Information revealed during interviews that provided further insight into specific components of a survey response were included in the results section of this study. Themes consistent across most interviews were used to inform the discussion section of this manuscript.

### Evaluation of publicly available information

To consider experiences of laboratories who conducted SARS-CoV-2 testing but did not participate in this study, public information was gathered to contextualize or provide further insight for survey and interview results. Publicly available information used for this analysis included peer-reviewed publications, online local or national news articles, white papers, and articles published by campus news. Google, Google Scholar, and the EBSCO database were used in these searches. Sample search terms included: (“SARS-CoV-2” OR “COVID19” OR “COVID-19”) AND (“veterinary diagnostic” OR “animal diagnostic” OR “veterinary lab” OR “animal lab”) AND (“university” OR “college” OR “school”). In some cases, when gathering information for specific laboratories, the name of the institution of interest was included in the search terms. Information gathered from these searches supplemented information presented in surveys or interviews.

## Results

In total, 76 invitations for participation were sent and 38 total responses were recorded, yielding a response rate of 50%. All regions of the US were accounted for by respondents, with most responding laboratories located in the Midwest and South (Fig 1). Thirty-three total states were represented (Fig 1).

**Fig 1.**
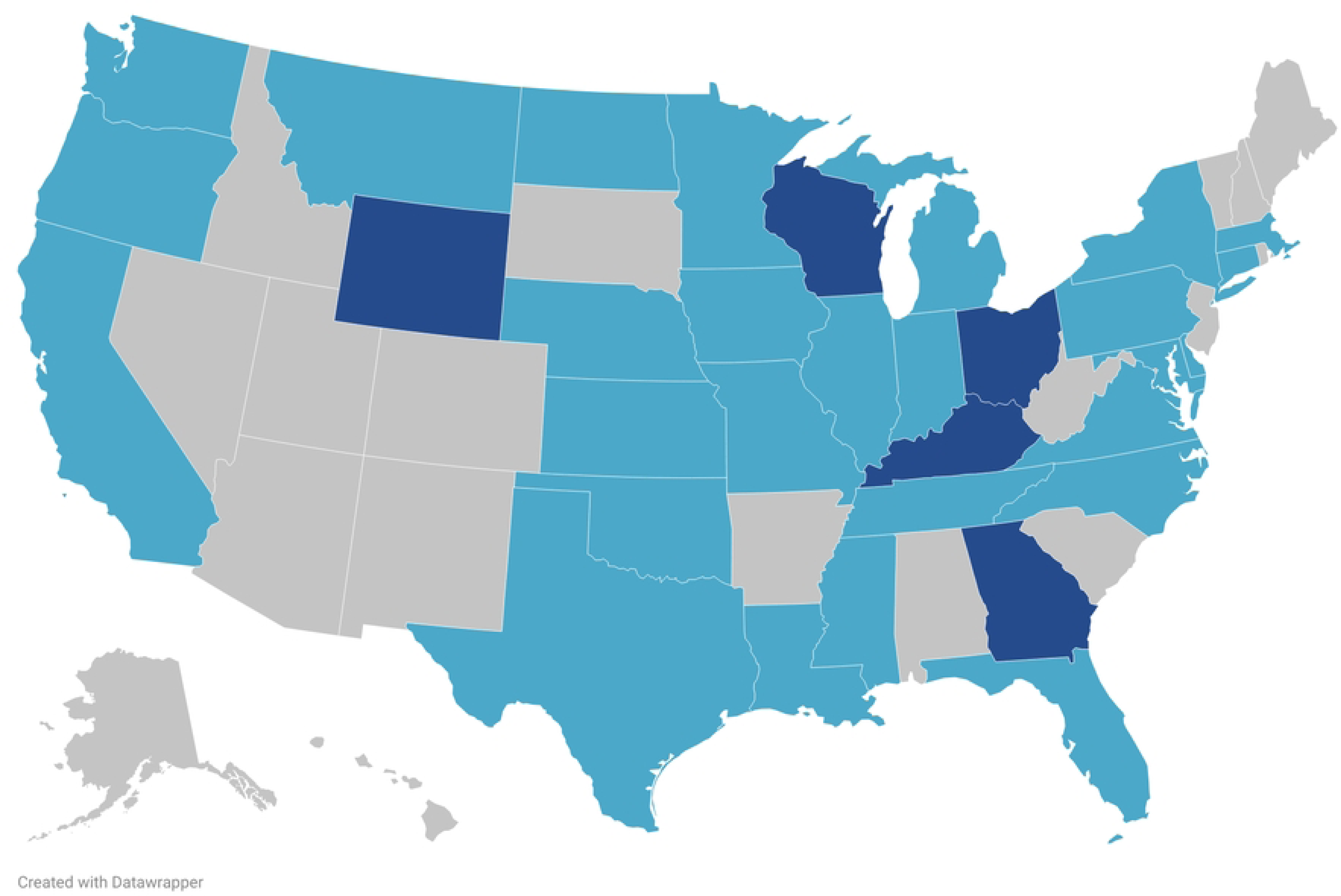
Map of Survey Respondents. Light blue represents one responding laboratory and dark blue represents two responding laboratories. Visualization made using Datawrapper.

Thirty (79%) of 38 responding veterinary diagnostic laboratories were operated within a university, of which 19 were operated within a college of veterinary medicine (Table 1). Eight laboratories (21%) were not affiliated with a college or university (Table 1). Of the 38 survey respondents, 20 (53%) conducted both animal and human SARS-CoV-2 testing, 7 conducted only animal testing, 4 conducted only human testing and 7 did not conduct human or animal tests (Table 1). More respondents conducted only animal testing than only human SARS-CoV-2 testing (7 laboratories versus 4 laboratories, respectively; Table 1).

**Table 1.** Organizational Structure and Distribution of Animal and Human SARS-CoV-2 Testing Among Responding Laboratories (n=38).

| **Organizational structure of laboratory** | **Number of laboratories (%)** |
| --- | --- |
| Operating within a college of veterinary medicine, partially/fully state funded | 6 (16) |
| Operating within a college of veterinary medicine, not state funded | 13 (34) |
| Operating within a university, but not a college of veterinary medicine, partially/fully state funded | 5 (13) |
| Operated within a university, but not a college of veterinary medicine, not state funded | 6 (16) |
| Not within a university or college of veterinary medicine, state funded | 7 (18) |
| Not within a university or college of veterinary medicine, federally funded | 1 (3) |
| **Testing conducted** |  |
| Conducted both animal and human SARS-CoV-2 testing | 20 (53) |
| Conducted animal SARS-CoV-2 testing but not human SARS-CoV-2 testing | 7 (18) |
| Conducted human SARS-CoV-2 testing but not animal SARS-CoV-2 testing | 4 (11) |
| Did not conduct animal or human SARS-CoV-2 testing | 7 (18) |

### Animal SARS-CoV-2 testing

Most laboratories that conducted animal SARS-CoV-2 testing began testing in early 2020 (58%), followed by mid-2020 (29%) and late 2020 (13%; n = 24). Furthermore, 23 (88%) surveyed laboratories were continuing to test animals as of May 2023 (n = 26).

The species of tested animals can be broadly categorized as wildlife (e.g., moose, bobcat), companion animals (e.g., cats, dogs), agricultural or farming animals (e.g., cattle, pigs), and zoo animals (e.g., tigers, lions). Among survey respondents, the animal category most tested was companion animals (42%), followed by wildlife and zoo animals (23% respectively; Fig 2). Farm animals were the least tested animal category (13%; Fig 2). These results are consistent with the United States Department of Agriculture reporting that companion animals were the main species tested, followed by wildlife and zoo animals.[15]

**Fig 2.**
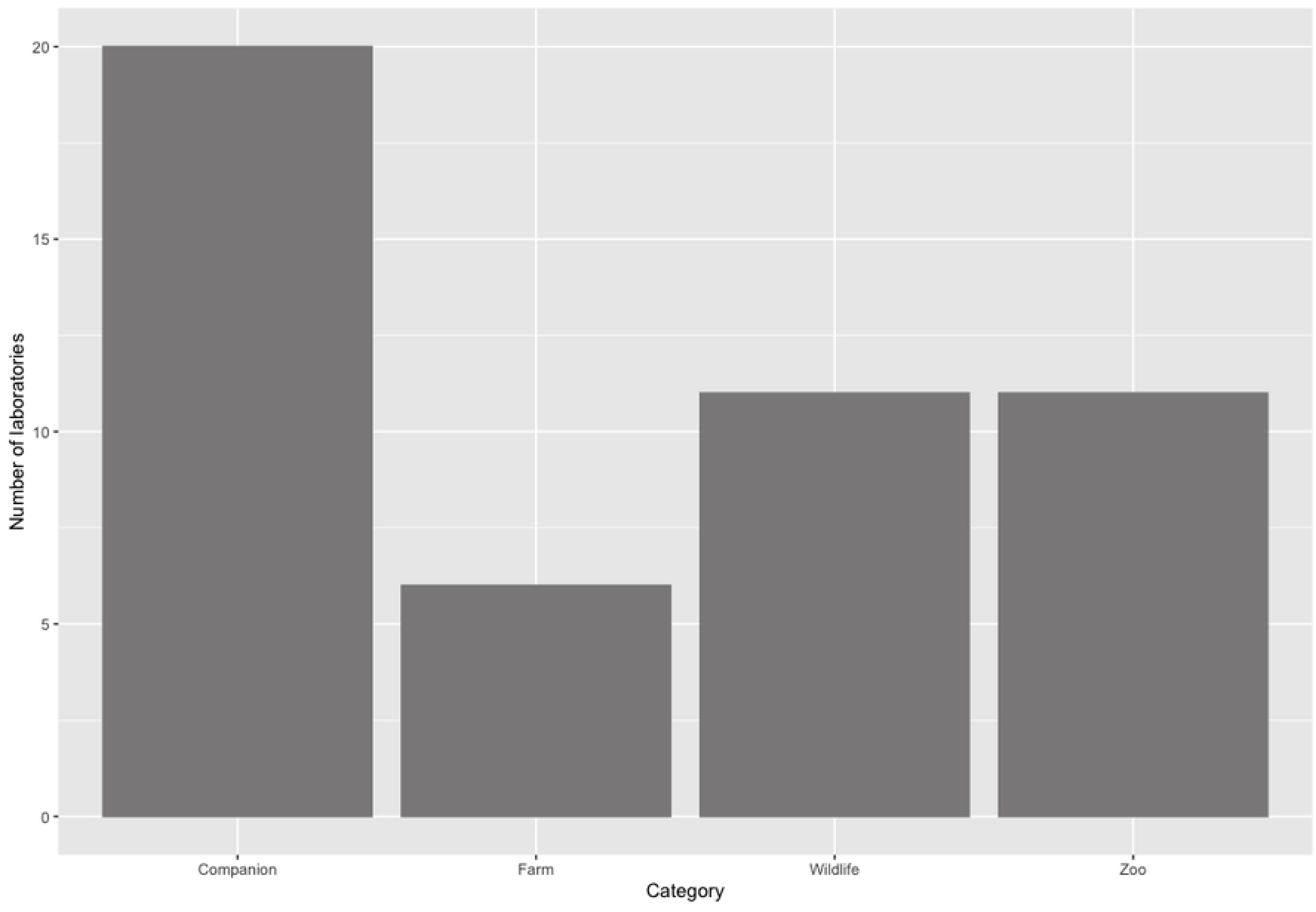
Frequency Distribution for Laboratories Testing Various Animal Species for SARS-CoV-2 Among Survey Respondents. Animal species were categorized into companion, farm, wildlife, and zoo animals and the bars represent the number of laboratories that tested for each category.

The number of animal samples tested for SARS-CoV-2 among 22 survey respondents who provided these numbers ranged from 9 to 3,600 animals, although this large range may be accounted for by some survey respondents including research tests in addition to clinical or diagnostic tests. Furthermore, it is possible that survey respondents reported the number of SARS-CoV-2 tests performed as opposed to the number of animals tested as some animals were tested serially. Every laboratory that conducted animal SARS-CoV-2 testing used reverse transcription polymerase chain reaction (RT-PCR) methodology for tests. In addition to RT-PCR testing, seven respondents (29%) performed viral nucleic acid sequencing to gather genotypic information about the viral variants.

### Human SARS-CoV-2 testing

Of the 38 laboratories providing survey responses, 24 laboratories (63%) indicated that they conducted human SARS-CoV-2 testing. Among these 24 laboratories, 14 (58%) conducted human SARS-CoV-2 testing for a university population and 7 (29%) tested for the general community. Three laboratories tested for both a university population and the general community (13% respectively). Two laboratories conducted testing for local hospitals (8%). Fourteen of the 17 universities whose laboratories conducted testing for their populations had testing requirements that ranged from compulsory testing among specific student populations or under certain conditions (e.g., student athletes, symptomatic individuals only) to the entire population (e.g. students, faculty and staff) requiring testing.

The most common reason for human sample testing was to increase university testing capacity (17 laboratories; 71%), followed by increasing testing capacity for the local community (6 laboratories; 25%) and testing to decrease turnaround time (5 laboratories; 21%). Furthermore, 4 laboratories tested to increase testing capacity for the local health departments and hospitals (17%) or to provide information to local health departments through sequencing (4 laboratories, 17%). Research and reducing test cost were also cited as a testing motivation.

As was the case for animal tests, real-time RT-PCR was the most common method of SARS-CoV-2 testing and used in 22 (92%) laboratories. In addition to RT-PCR, laboratories also conducted enzyme-linked immunosorbent assays and sequencing. Most real-time RT-PCR tests targeted ORF1ab, spike, and nucleocapsid genes. As displayed in Fig 3, nasopharyngeal swabs were the most common sample type (14 laboratories; 58%) followed by anterior nares swabs (12 laboratories; 50%) and saliva (10 laboratories; 42%). Only one institution collected samples from nasal mid-turbinate swabs (4%) and oropharyngeal swabs (4%).

**Fig 3.**
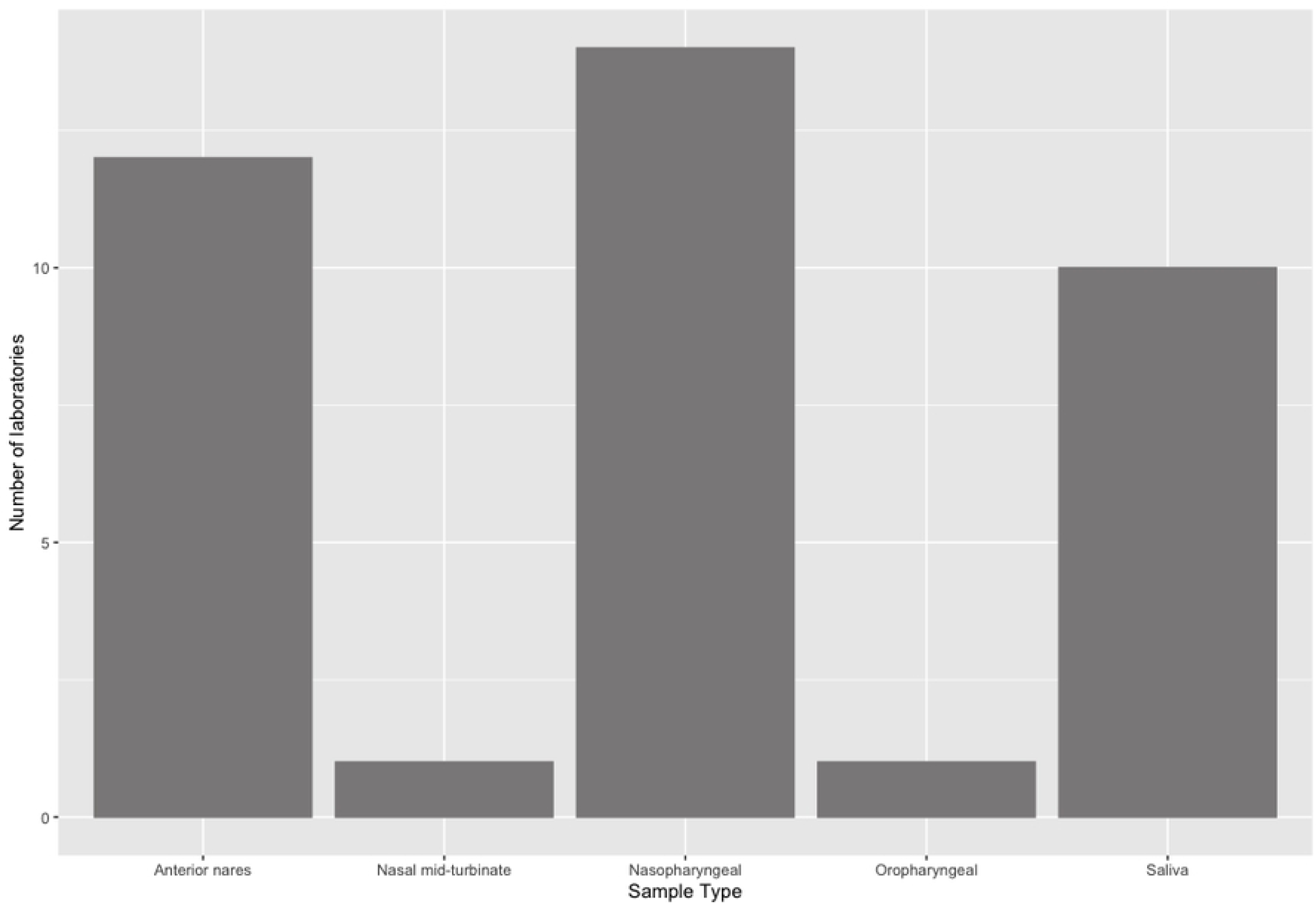
Frequency Distribution of the Sample Types Tested for SARS-CoV-2 Among the 24 Laboratories Surveyed That Conducted Human Testing.

Only 1 institution of the 24 that conducted human SARS-CoV-2 testing was responsible for sample collection. At this institution, observed self-collection sampling occurred at designated campus testing sites. Furthermore, one institution pooled diagnostic test samples and two laboratories pooled surveillance tests. At the institution that pooled diagnostic tests, samples were pooled in groups of five. When pools were positive, individual samples were tested separately following deconvolution of the pools. Seventeen (74%) laboratories that tested human samples used automation or robotics to achieve their testing capacity with the Kingfisher Flex most frequently cited for high-throughput nucleic acid extraction. For human SARS-CoV-2 testing, 9 (39%) institutions conducted viral nucleic acid sequencing to gather genotypic information about the viral variants. Two of the fourteen laboratories that did not conduct viral nucleic acid sequencing sent samples to state collaborators for sequencing.

Most laboratories began their human SARS-CoV-2 testing program in the late summer months of 2020. Most of these laboratories ended their testing programs mid-way through 2022, though one ended as early as July 2020 and one was continuing testing until May 2023.

Maximum samples tested per day and total samples tested among the human SARS-CoV-2 testing laboratories varied greatly across laboratories (Table 2). Across all laboratories, daily testing capacity ranged from 300 to 14,000 samples tested per day, with a median of 2,000 samples per day. The total number of samples tested ranged from 6,215 to 3,000,000 total samples, with a median number of total samples tested of 121,546. There was no statistically significant relationship between samples tested per day and result turnaround time (Spearman correlation coefficient: -0.06; p = 0.789 > 0.05).

**Table 2.**
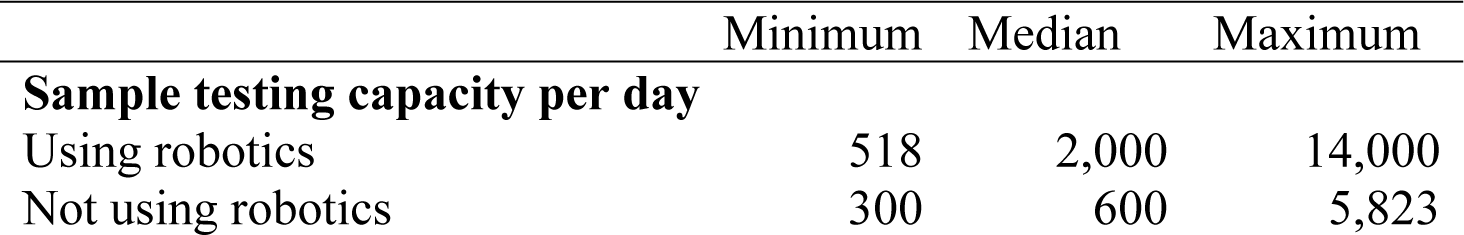
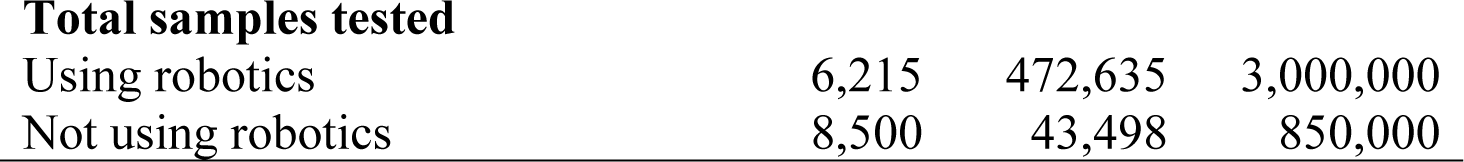
Summary Statistics of Daily Testing Capacity and Total Samples Tested Among Laboratories That Conducted Human SARS-CoV-2 Testing, Stratified by Laboratories that Used Robotics Versus Did Not Use Robotics (n = 24).

|  | Minimum | Median | Maximum |
| --- | --- | --- | --- |
| **Sample testing capacity per day** |  |  |  |
| Using robotics | 518 | 2,000 | 14,000 |
| Not using robotics | 300 | 600 | 5,823 |
| **Total samples tested** |  |  |  |
| Using robotics | 6,215 | 472,635 | 3,000,000 |
| Not using robotics | 8,500 | 43,498 | 850,000 |

### Automation and Pooling

Laboratories using robotics tested more samples per day and tested more samples total than laboratories that did not use robotics (Table 2). The median number of samples tested per day was 2,000 for laboratories that used robotics and 600 for laboratories that did not use robotics (Table 2). Similarly, the median total number of samples tested for laboratories that used robotics was 472,635 samples versus a median number of samples of 43,498 samples for laboratories that did not use robotics (Table 2).

While automation was used for laboratories that had higher testing capacities, automation was not *essential* to achieving a high testing capacity. One laboratory reported a daily testing capacity of 5,823 samples tested per day without use of robotics, which is substantially higher than the median daily testing capacity of 600 samples tested per day among the non-automated group. This laboratory achieved this high sample testing capacity by having a large workforce totaling approximately 90 full time equivalents. The laboratory tested one-third of the student body each week such that the entire study body was evaluated every three weeks, enabling the university to detect and address hotspots of SARS-CoV-2 cases earlier and ultimately reduce the burden of SARS-CoV-2 on campus.

One laboratory conducted pooling for diagnostic SARS-CoV-2 tests and, using this methodology, achieved a sample testing capacity of greater than 10,000 samples per day. Pooling was conducted in 96-welled plates, each containing 93 pools of 5 samples. Processed samples were placed into barcode-visible Biomek tube racks so that Biomek i5 could generate pools while also allowing the Data Acquisition and Reporting Tool 2.0 software to record the position of each sample in the pooled wells. The well position within the 96-well plate and the sample position in storage racks were retained and managed through a custom software application (COVID-19 Receiving App).[16] Three wells were left as controls.[16] In forty-five minutes, 465 samples were pooled into 93 wells of the 96-well plate.[16] If any pools tested positive, deconvolution took place and samples were tested individually.[16] Result reporting for pooled samples had a median turn-around time of 4 hours and 46 minutes, although positive pools had a median turn-around time of 22 hours and 19 minutes.[16] Samples that were not pooled (and tested individually) had a median turn-around time of 3 hours and 56 minutes.[16] Although logistically more complicated, pooling increased efficiency, enabling rapid test result communication, which ultimately improved the effectiveness of intervention among positive cases. Pooled testing also resulted in lower cost per sample tested due to lower workforce requirements and conservation of testing reagents and supplies.

Pooling, however, was not required to achieving a high sample testing capacity. The laboratory with the highest number of samples tested of all surveyed laboratories, with a daily sample testing capacity of 14,000 samples tested and 3,000,000 samples tested overall, did *not* pool samples for testing. This strategy, however, involved a large number of full time FTE’s and extended/double shifts.

While some laboratories did consider pooling, one barrier to using pooling in the diagnostic workflow included lack of access to the proper equipment to conduct pooling effectively. One laboratory noted that the use of saliva samples and the lack of access to liquid sensing tips caused the sample to dribble throughout the machine during pooling trials. Given the low population size on this institution’s campus during the pandemic, they decided to focus on testing optimization in other areas of the testing scheme instead of pooling. A different laboratory noted that pooling was not pursued because the deconvolution of positive samples would have increased testing cost and sample result turnaround times. Given these barriers, the decision to pool as a mechanism of increasing testing capacity was dependent on resources, such as workforce, lab supplies, and budget.

### Barriers to testing

Seventeen (71%) laboratories reported that obtaining the proper certification or licensure was an initial impediment to testing human SARS-CoV-2 samples (Table 3). The Clinical Laboratory Improvement Amendments (CLIA) certification was the most frequently cited barrier related to licensure for testing. Limited human workforce capacity and having other organizational responsibilities were also primary barriers to testing human samples (58% and 38% respectively; Table 3). Three laboratories cited lack of qualified workforce and the need for constant staff training to ensure consistent results as additional issues. These same laboratories also indicated that managing work schedules to ensure testing capacity while preventing burn out among employees was challenging. One institution stated that tripling the size of their workforce in one month to accomplish SARS-CoV-2 testing was easy, as graduate students, professors, and department heads took pride in volunteering for the testing initiative. Given that normal operations were interrupted, many students and faculty had more time to dedicate towards a testing initiative, requiring this institution to limit the number of volunteers working in the diagnostic laboratory. Limited physical space for testing and obtaining funding were the least frequently reported barriers to human testing (21% and 17% respectively), and none of the surveyed laboratories reported achieving satisfactory testing accuracy as a barrier to testing operations (Table 3).

**Table 3.** Cited Barriers to Human SARS-CoV-2 Testing Operations Among Respondents (n = 24).

| Barrier | Frequency (%) |
| --- | --- |
| Obtaining the proper certifications and/or licensure | 17 (71) |
| Limited human workforce capacity | 14 (58) |
| Other organizational responsibilities | 9 (38) |
| Limited testing equipment, materials, and supplies | 8 (33) |
| Other (e.g., billing insurance, IT issues, state or local leadership) | 8 (33) |
| Difficulty coordinating with other organizations | 7 (29) |
| Limited physical space for testing | 5 (21) |
| Obtaining funding | 4 (17) |
| Achieving satisfactory test accuracy | 0 (0) |

One frequently cited barrier to SARS-CoV-2 testing was institutional leadership. Four laboratories noted that university administration constrained SARS-CoV-2 testing initiatives. In one case, administration viewed SARS-CoV-2 as not sufficiently urgent to warrant resources to mount a testing response. For this academic institution—and another institution that cited internal leadership problems as a barrier to operation—there was an additional concern that reporting SARS-CoV-2 numbers on campus would result in diminished enrollment in the universities. To justify the need for SARS-CoV-2 testing despite these concerns, one institution reported that resources were allotted to testing once a county public health agency became involved to advocate for the importance of a testing program.

Support by human health agencies (e.g., public health agencies, human health providers) for veterinary diagnostic laboratories to participate in SARS-CoV-2 response was not universal, as two laboratories cited human health agencies as a barrier to developing a testing program. One institution described how public health groups made the state’s granting of CLIA certification difficult for the veterinary diagnostic laboratory. In this case, the institution reported that these human health agencies eventually realized the positive impact that the veterinary laboratories could have on achieving the testing capacity required to control the pandemic. Consequently, the veterinary diagnostic laboratory began surveillance testing, resulting in a surge of the state’s SARS-CoV-2 positivity rate, and then consequently the granting of CLIA licensure for the laboratory to begin diagnostic testing. While this ultimate approval for diagnostic testing enabled a higher testing capacity to be achieved in this state, this response was delayed.

## Discussion

Many veterinary diagnostic laboratories developed successful programs for testing human samples for SARS-CoV-2, applying high throughput and high complexity diagnostic principles used in their routine testing operations. A total of 8,273,602 human samples were tested by surveyed laboratories participating in this study. One university’s veterinary diagnostic laboratory tested one-third of their university students each week to ensure that every student was tested every three weeks, allowing for the identification of outbreak clusters on campus and timely intervention by public health groups. Another university, early in the pandemic, determined that the student body would need to be tested twice per week to allow faculty, staff and students to safely return to work and school, requiring a testing capacity of 7,000 samples tested per day.[17,18] This animal diagnostic laboratory, using its expertise and population testing principles and workflows established a final daily testing capacity exceeding 10,000 samples with a result turnaround time of 24 to 36 hours, ultimately testing 2,079,685 samples in a rural region where closure of the university would have had large impacts on accessibility of testing for local community members. Some veterinary diagnostic laboratories offered testing services to nearby institutions and satellite campuses and consequently, some of these university testing programs, with their animal population based high throughput design, made up a large proportion of total state tests. One laboratory’s testing capacity of 18,000 samples daily corresponded to 20% of all daily testing completed in that state, and to 1.5-2.5% of all SARS-CoV-2 daily samples tested nationwide.[19] One other veterinary diagnostic laboratory conducted 25% of its state’s total tests; In fact, this laboratory tested four times as many SARS-CoV-2 samples than the state’s public health laboratories[12] and more than any other laboratory in the state. Given these contributions, veterinary diagnostic laboratories have demonstrated their importance and positive impact in achieving high testing capacity on population-wide scale during public health emergencies.

These contributions to states’ large scale testing programs were achieved despite significant challenges, with the most frequently cited obstacle being the regulatory requirements that prevented veterinary diagnostic laboratories from participating either early in the pandemic or, depending on states’ interpretation of rules and regulations, throughout the pandemic.

There were two testing options for SARS-CoV-2 laboratories: surveillance testing and diagnostic testing. The Food and Drug Administration authorized surveillance testing for emergency use during the SARS-CoV-2 pandemic, allowing laboratories—including veterinary diagnostic laboratories—to monitor SARS-CoV-2 incidence in populations.[20] Surveillance testing, however, does not permit contact tracing of detected cases (i.e. tracing back results to tested individuals and determining potential infected and infectious contacts), thereby decreasing the public health usefulness of surveillance testing programs.[20] To conduct diagnostic testing which allows identification of the infected patient and subsequent contact tracing, laboratories must obtain Clinical Laboratory Improvement Amendments (CLIA) certification with requirements that were difficult, or depending on states’ interpretation, impossible to meet for veterinary diagnostic laboratories. The requirement in 42 CFR §493.1443 in particular states that a laboratory director must be a licensed medical professionals (i.e., Doctor of Osteopathic Medicine, Medical Doctor, Doctor of Veterinary Medicine) with training and professional experience limited to human or public health diagnostic testing, specifically that “[l]aboratory testing of non-human specimens is not acceptable experience, e.g., environmental, animal testing.”[21] This requirement prevented some veterinary diagnostic laboratories from participating at all despite their expertise, experience and capacity in testing more samples with “the same machinery, scientific theory, and tests as human health facilities with the only difference being the taxonomy of the sample source.”[12] A case in point was that veterinary laboratory diagnosticians in one state were called upon to train human health professionals to perform the testing that they themselves were not certified to perform and participate in the SARS-CoV-2 testing response. Clinical Laboratory Improvement Amendments (CLIA) certification thus constituted a “bureaucratic wall” in many states, requiring the establishment of sometimes complex partnerships of veterinary diagnostic laboratories with state or private human health partners.[22] Two interviewed laboratories obtained rapid and independent certification, due to support within established networks in their State Department of Health, or due to state legislators’ support, as well as strong advocacy by university leadership.

Most animal diagnostic laboratories, however, faced significant barriers in obtaining CLIA certification due to challenges in contracting with a human health partner and strict state interpretations of CLIA requirements. Differences in CLIA enforcement state-to-state certainly contributed to inequities in testing resources across the country. Attitude towards diagnostic testing by veterinary laboratory also constituted an obstacle in some states, despite veterinary and human diagnostic laboratories using the same diagnostic methods. In fact, restrictions on one veterinary diagnostic laboratory to only provide surveillance testing with confirmation of positive findings at the public health laboratory were lifted within days of program commencement, resulting in lifted restrictions only once the public health laboratory could not cope with the testing demand.

There is an urgent need when preparing for public health emergencies to reduce barriers and consider available high quality testing capacity in veterinary diagnostic laboratories. Veterinary diagnostic laboratories, often located in rural areas, can have a significant impact on increasing accessibility to testing among rural residents. For example, the state of Washington had fifteen operating SARS-CoV-2 testing laboratories. Fourteen of these laboratories were in the state’s Western region, with the one animal diagnostic laboratory in the East filling an important gap in accessibility to testing.

Despite the important role that veterinary diagnostic laboratories played in the pandemic response, there is insufficient support from non-animal health institutions, ultimately impeding animal diagnostic laboratory’s abilities to achieve CLIA certifications. One interviewee noted:

> “We, diagnostic veterinary labs, continue to see push back from public health laboratories. The way we function, the qualifications, are equivalent to CLIA lab directors but because of legislation we can’t just jump in and do certain things. The best outcome at a national level to this effort and for preparedness for the future would be for vet labs to be at least permitted on a stand-by basis to jump in without hurdles from CLIA.”

This interviewee also describes how the “nation is losing out on capacity by excluding the possibility that in an emergency basis, that [animal diagnostic laboratories] could jump in and get going.” The SARS-CoV-2 pandemic helped convey the impact that veterinary animal laboratories may have in population health emergencies despite the presence of significant barriers, indicating a need for change.

Unfortunately, despite this need for change, on July 26, 2022, the Centers of Medicare & Medicaid Services (CMS) proposed revisions to CLIA in the *Federal Register, Vol. 87, No. 142* stating that they “believe that DVM should be removed from the qualifying doctoral degrees as it is not relevant to testing on specimens derived from the human body” despite “CMS recogniz[ing] that the COVID-19 public health emergency (PHE) requires flexibility, and [they are] are committed to taking critical steps to ensure America’s clinical laboratories can respond during a PHE to provide reliable testing while ensuring patient health and safety.” [9] The American Association of Veterinary Laboratory Diagnosticians responded to this proposed revision by stating[9]:

> “The incorporation of [testing by veterinary diagnostic laboratories] into the [COVID-19 public health emergency] response was often significantly delayed due to the inflexibility regarding recognition of [veterinary diagnostic laboratory] staff’s training, knowledge and experience as being sufficiently equal to that mandated under CLIA. It is paradoxical that on the heels of the strong participation [of veterinary diagnostic institutes] in the COVID-19 [public health emergency], the new proposal actually seeks to further restrict any recognition of veterinary training and degrees as being germane to the oversight of diagnostic testing of human samples.”

Given the undeniable and positive impact of veterinary diagnostic laboratory participation in the COVID-19 response, policymakers and regulators should focus on reducing regulatory impediments to enhance the Nation’s preparedness and emergency response capacity.

## Conclusion

Veterinary diagnostic laboratories were major contributors to the Nation’s COVID-19 pandemic public health response by adding to local and regional SARS-CoV-2 testing capacity and assisting in “flattening the epidemic curve.” It is imperative that human health agencies and federal regulators recognize these contributions and readily collaborate with and call on the expertise and resources of the veterinary community and diagnostic centers as new public health threats emerge. More than 60% of human infectious diseases have a zoonotic origin, which underscores the urgent need to promote animal and human diagnostic collaboration under the One Health umbrella in preparation for future pandemics.[23]

## Data Availability

Data cannot be shared publicly because data is not anonymous. The data includes sensitive information regarding specific institutions and their experiences during COVID-19. The corresponding author will provide data upon request.

https://cornell.box.com/s/bssw7jmadyrckxd4ahj3yy2if3u3e7nk

## Supporting Information

**S1 File. Survey Distributed to Veterinary Diagnostic Laboratories.** Following formal survey approval by the American Association of Veterinary Medical Colleges, this online cross-sectional Qualtrics survey was distributed to American Association of Veterinary Medical Colleges and the National Animal Health Laboratory Network member laboratories in the United States.

**S2 File. Interview Questions.** To learn more about the specific elements of a laboratory’s response, a convenience sample of survey respondents were asked to participate in a semi-structured interview.

## References

1. Chams N, Chams S, Badran R, Shams A, Araji A, Raad M, et al. COVID-19: A Multidisciplinary Review. Front Public Health [Internet]. 2020 [cited 2023 Feb 27];8. Available from: https://www.frontiersin.org/articles/10.3389/fpubh.2020.00383

2. Patel A. Initial Public Health Response and Interim Clinical Guidance for the 2019 Novel Coronavirus Outbreak — United States, December 31, 2019–February 4, 2020. MMWR Morb Mortal Wkly Rep [Internet]. 2020 [cited 2023 Feb 27];69. Available from: https://www.cdc.gov/mmwr/volumes/69/wr/mm6905e1.htm

3. Sanche S, Lin YT, Xu C, Romero-Severson E, Hengartner N, Ke R. High Contagiousness and Rapid Spread of Severe Acute Respiratory Syndrome Coronavirus 2 - Volume 26, Number 7—July 2020 - Emerging Infectious Diseases journal - CDC. [cited 2023 Mar 8]; Available from: https://www.nc.cdc.gov/eid/article/26/7/20-0282_article

4. Fenemigho I, Ukponmwan E, Nnakwue EC, Udoete I, Asuzu C, Adaralegbe A, et al. COVID-19, flattening the curve: recommendations towards control and managing a second wave. J Glob Health Rep. 2020 Aug 10;4:e2020074.

5. New COVID-19 Cases Worldwide [Internet]. Johns Hopkins Coronavirus Resource Center. [cited 2023 Mar 8]. Available from: https://coronavirus.jhu.edu/data/new-cases

6. Population, total - Korea, Rep. | Data [Internet]. The World Bank. [cited 2023 Mar 15]. Available from: https://data.worldbank.org/indicator/SP.POP.TOTL?end=2021&locations=KR&start=1960

7. Issac A, Stephen S, Jacob J, VR V, Radhakrishnan RV, Krishnan N, et al. The Pandemic League of COVID-19: Korea Versus the United States, With Lessons for the Entire World. J Prev Med Pub Health. 2020 Jul;53(4):228–32.

8. World Organisation for Animal Health. Veterinary Laboratory Support to the Public Health Response [Internet]. 2020 [cited 2023 Mar 14]. Available from: https://rr-americas.woah.org/en/news/veterinary-laboratory-support-to-the-public-health-response/

9. Jerry Saliki. (American Association of Veterinary Laboratory Diagnosticians). Letter to: Undisclosed recipients. 2022 Sept 22.

10. Janet Donlin. (American Veterinary Medical Association). Letter to: Sarah Bennett and Cindy Flacks (Centers for Medicare & Medicaid Services). 2022 Sept 23.

11. Taylor LH, Latham SM, Woolhouse ME. Risk factors for human disease emergence. Philos Trans R Soc B Biol Sci. 2001 Jul 29;356(1411):983–9.

12. This veterinary lab is the linchpin in one state’s coronavirus testing approach - The Washington Post [Internet]. [cited 2023 Sep 11]. Available from: https://www.washingtonpost.com/science/2020/05/12/this-veterinary-lab-is-linchpin-one-states-covid-19-testing-approach/

13. Cima G. Animal health laboratories aid testing for COVID-19 in people [Internet]. American Veterinary Medical Association. [cited 2023 Mar 14]. Available from: https://www.avma.org/javma-news/2020-06-01/animal-health-laboratories-aid-testing-covid-19-people

14. Nolen RS. Veterinary labs continue to support COVID-19 testing [Internet]. American Veterinary Medical Association. [cited 2023 Mar 14]. Available from: https://www.avma.org/javma-news/2020-07-01/veterinary-labs-continue-support-covid-19-testing

15. USDA APHIS | Cases of SARS-CoV-2 in Animals in the United States [Internet]. [cited 2023 Sep 11]. Available from: https://www.aphis.usda.gov/aphis/dashboards/tableau/sars-dashboard

16. Laverack M, Tallmadge RL, Venugopalan R, Sheehan D, Ross S, Rustamov R, et al. The Cornell COVID-19 Testing Laboratory: A Model to High-Capacity Testing Hubs for Infectious Disease Emergency Response and Preparedness. Viruses. 2023 Jul;15(7):1555.

17. Ramanujan K. Cornell teams work tirelessly to limit COVID spread [Internet]. Cornell Chronicle. [cited 2023 Mar 14]. Available from: https://news.cornell.edu/stories/2020/10/cornell-teams-work-tirelessly-limit-covid-spread

18. Frazier PI, Cashore JM, Duan N, Henderson SG, Janmohamed A, Liu B, et al. Modeling for COVID-19 college reopening decisions: Cornell, a case study. Proc Natl Acad Sci U S A. 2022 Jan 11;119(2):e2112532119.

19. U Of I Lab Working To Reduce Time To Get Results Of Saliva COVID-19 Tests [Internet]. NPR Illinois. 2020 [cited 2023 Sep 11]. Available from: https://www.nprillinois.org/illinois/2020-09-03/u-of-i-lab-working-to-reduce-time-to-get-results-of-saliva-covid-19-tests

20. Health C for D and R. FAQs on Testing for SARS-CoV-2. FDA [Internet]. 2023 Aug 16 [cited 2023 Sep 12]; Available from: https://www.fda.gov/medical-devices/coronavirus-covid-19-and-medical-devices/faqs-testing-sars-cov-2

21. 42 CFR 493.1443 -- Standard; Laboratory director qualifications. [Internet]. [cited 2023 Sep 11]. Available from: https://www.ecfr.gov/current/title-42/part-493/section-493.1443

22. Tewari D, Zeman DH. 2020, a year to remember: AAVLD perspective. J Vet Diagn Investig Off Publ Am Assoc Vet Lab Diagn Inc. 2021 May;33(3):393–5.

23. Weiss RA, Sankaran N. Emergence of epidemic diseases: zoonoses and other origins. Fac Rev. 2022 Jan 18;11:2.

